# City Reduced Probability of Infection (CityRPI) for Indoor Airborne Transmission of SARS-CoV-2 and Urban Building Energy Impacts

**DOI:** 10.1101/2021.01.19.21250046

**Authors:** Ali Katal, Maher Albettar, Liangzhu (Leon) Wang

## Abstract

Airborne transmission of aerosols produced by asymptomatic individuals is a large portion of the SARS-CoV-2 spread indoors. Outdoor air ventilation rate, air filtration, room occupancy, exposure time, and mask-wearing are among the key parameters that affect its airborne transmission in indoor spaces. In this work, we developed a new web-based platform, City Reduced Probability of Infection - CityRPI, to calculate the indoor airborne transmission of COVID-19 in various buildings of a city scale. An archetype library of twenty-nine building types is developed based on several standards and references. Among the mitigation strategies recommended to reduce infection risk, some could result in significant energy impacts on buildings. To study the combined effects of energy consumption and reduced infection probability, we integrated CityRPI with City Building Energy Model. We applied the integrated model to Montreal City and studied the impact of six mitigation measures on the infection risk and peak energy demand in winter. It shows that the same strategy could perform quite differently, depending on building types and properties. In the winter season, increasing the outdoor air ventilation rate may cause massive building energy consumption. All strategies are shown to reduce the infection risk but wearing a mask and reducing exposure time are the most effective strategies in many buildings, with around 60% reduction. Doubling the outdoor air ventilation rate is not as effective as other strategies to reduce the risk with less than 35% reduction. It also significantly increases building peak heating demand with 10-60%.

## 1. Introduction

A novel coronavirus disease, later named SARS-CoV-2 or COVID-19, rapidly spread throughout the world and was announced as a pandemic on March 11, 2020 [1]. As of December 25, 2020, the total number of COVID-19 cases exceeded 78.1 million worldwide, with a confirmed 1,736,752 total deaths [2]. The main routes of transmission of SARS-CoV-2 are direct and indirect contact transmission with the infected people, respiratory transmission by large droplets within 1 m distance from an infected person, and transmission by airborne aerosols [3–5]. Human expiratory activities such as coughing, sneezing, speaking, singing, and even breathing release particles in a wide range of sizes, with most of them between 2 and 100 *μm* [6]. The small respirable particles < 5 − 10 *μm* can remain airborne and can spread at long distances. The largest droplets fall next to a source, within a distance of 1–2 m, as a result of gravitational force [7]. The latest research findings show that aerosols’ airborne transmission is a large portion of the spread of COVID-19 disease, especially in indoor spaces with poor ventilation conditions, large gathering, and long-duration exposure to high concentrations of aerosols [7–11]. Several studies show that improving the ventilation condition, wearing a face mask, avoid overcrowding, and shortening the event time (exposure time) can significantly reduce the airborne infection risk in indoor environments [12–14]. Buonanno et al. (2020b) evaluated the number of people infected by asymptomatic SARS-CoV-2 subjects in indoor environments. The results show the great importance of proper ventilation in the containment of the virus in indoor environments. Curtius et al. (2020) tested the efficiency of operating four air purifiers equipped with HEPA filters and the total air exchange rate of 5.5 *h*^-1^ in a high school classroom. The aerosol concentration was reduced by more than 90 % within less than 30 minutes after running the purifiers. Some other researchers evaluated the reduced infection risk using other strategies. For example, Dai and Zhao (2020) calculated the required ventilation rate to ensure a probability of less than 1% infection for different exposure times using the Wells-Riley model [18]. They modeled some typical scenarios and concluded that the minimum required ventilation rate can be reduced to a quarter by wearing the mask, which can be achieved by normal ventilation mode. Lelieveld et al. (2020) estimated the infection risk in several indoor environments and compared the infection risk between different scenarios. Active room ventilation and the wearing of face masks by all subjects could reduce the individual infection risk by five to ten, similar to a high-volume HEPA air purifier. Zhang (2020) calculated the reduced infection risk by integrating different air quality strategies, including source control, ventilation, and air cleaning strategies. They modeled classrooms and offices because of their large occupant density. Integration of strategies can reduce the infection risk by a factor of 8.5 to 500.

Miller et al. (2020) studied a super-spreading event at a weekly rehearsal in Skagit Valley, Washington, USA. Among the 61 attendees at the rehearsal, 53 cases were subsequently identified as infected. They used a modified Wells-Riley formulation [21] to calculate the airborne infection risk of COVID-19. The results highlighted four essential factors that increase the risk of aerosol transmission indoors: dense occupancy, long duration, loud vocalization, and poor ventilation. Sun and Zhai (2020) modified the Wells-Riley model by introducing two indices for social distancing and ventilation effectiveness. It was shown that half occupancy density could reduce the infection risk by 20-40% in the first 30 minutes of the event. The combination of proper social distance and high ventilation effectiveness can reduce the required ventilation rate for a safe indoor environment. Jiminez et al. (2020) extended Miller’s work to calculate the airborne infection risk in indoor spaces based on the disease’s prevalence rate in the area of study. Their model also provides much information on key input parameters based on the most recent COVID-19 studies and makes it possible to evaluate infection risks and mitigation strategies through a public data spreadsheet known as “COVID-19 Aerosole Transmission Estimator”.

Some researchers worked on the community scale reduction strategies of airborne transmitted diseases. Gao et al. (2009) developed a model for calculating indoor airborne infection risk at a community level and estimated the impact of control strategies such as ventilation rate on the infection risk. They concluded that the current ventilation rate in Hong Kong offices and the recommended value by ASHRAE be too low to control airborne infectious diseases in indoor environments. Gao et al. (2016) designed an indoor transmission network model for an ideal city to estimate the infection risk and disease spread in their other work. They estimated the effectiveness of using ventilation strategy in controlling the diseases spread and compared it with other intervention strategies. They concluded that the ventilation rate in homes, classrooms, and offices could reach up to 5 ACH by opening the windows. Its impact is the same as isolating 12% of the population or mass vaccination. They did not model all buildings in a city and used some actual censuses, social behavior surveys, building surveys, etc., for community-scale analysis.

The existing studies, e.g., Jiménez et al.’s estimator, evaluated the mitigation measures of reducing airborne transmission risk in limited buildings or specific cases, e.g., choir, classroom, subway, supermarket, and stadium. In contrast, the effectiveness of mitigation measures can vary with building type because of different ventilation conditions, occupants’ density, event time, dominant age of occupants, etc. Therefore, it is essential to cover different building types, preferably at an urban scale for multiple cities, and investigate the impact of different strategies in reducing airborne infection risk based on the building usages. Each city or state also has its unique COVID-19 prevalence with daily variations; e.g., the top five most infected state prevalence rate varies from 3.5%-5% in the U.S. and 0.6%-1.3% in Canada on December 29, 2021 [25]. Therefore, a city-based infection risk estimator is expected to connect its prevalence rate to daily infection risk directly.

Besides the variations of the prevalence rates, the infection risk model often has other key input parameters, often related to cities, buildings, and occupancies, e.g., lockdown dates and rates, occupancy levels, age, sex, and exposure time. Building system-specific parameters are also important, including floor area or room size, outdoor air ventilation, recirculation rates, duct filter types, with/without air cleaners and their capacities, and mask types. The acquisition of all this information at the urban scale may be achieved in two ways. One is to develop an archetype building library based on the publicly available data, such as building standards and codes and statistics database. Because an archetype building is specific to a building type, the results of urban-scale infection risk and mitigation strategies can be compared statistically among various building types and total numbers in a city. Although an actual building’s information may deviate from the archetype-based database, it is reasonable to consider that most buildings of the same type tend to follow similar standards and codes, and statistics. So the infection risk and mitigation evaluations are substantiated at the urban scale. A similar method has been widely applied to urban-level building energy analysis: Archetype libraries are developed to characterize the properties necessary for buildings’ energy analysis, but it is challenging to collect them for all buildings in a city.

On the other hand, when a particular building needs to be assessed, much of the required information is private and better provided by the building owner or a person who has access to the data. This may be achieved by a user-friendly interface, preferably based on a web service, so that a layperson without any knowledge about infection calculation can provide the information required and assess the infection risk and evaluate the mitigation strategies specific to his/her building.

In this study, we tried to achieve both solutions. First, we developed an archetype library based on different standards and codes to estimate the parameters we need to calculate infection risk in buildings. For example, a building’s minimum required ventilation rate is based on ASHRAE Standard 62.1 [26]. This work aims to evaluate the probability of infection in buildings based on the current standards and then reduce the infection risk by comparing different mitigation strategies. Second, we developed a user-friendly web-based and 3D-city portal, the City Reduced Probability of Infection for Indoor Airborne Transmission of COVID-19 (https://concordia-cityrpi.web.app/) for North America. The website facilitates any layperson to use the service without prior technical knowledge from his/her mobile devices or computers. All the archetype library data are based on publicly available data. All the private input parameters are not collected and discarded at the client-side after an active web session.

Moreover, many risk mitigation strategies create significant building energy impacts: e.g., increasing outdoor ventilation rates to 130% or 200% of the baseline during the winter of 2020 may increase the building’s total energy consumption and peak demand. It could overload the existing system and result in a power outage at an urban or community scale. Therefore, while reducing the probabilities of indoor infections is essential, it is equally important to evaluate each mitigation strategy’s energy impact at both a building and an urban scale to be implemented.

Therefore, we integrated CityRPI, which calculates the airborne infection risk of COVID-19 in all buildings, with CityBEM (City Building Energy Model) [27,28], which calculates the impact of different strategies on the buildings’ peak energy demand. In this paper, we performed the simulation for the coldest period of Winter 2019 in the City of Montreal, Canada. The weather data are obtained from the High-Resolution Deterministic Prediction System (HRDPS) developed by Environment and Climate Changa Canada (ECCC) [29]. The integrated model could be used for short-term forecasting of infection risk and energy consumption of buildings in the Winter of 2020.

## 2. Methodology

Figure 1 shows the integrated CityRPI and CityBEM model explained in detail in the following sections.

**Fig. 1.**
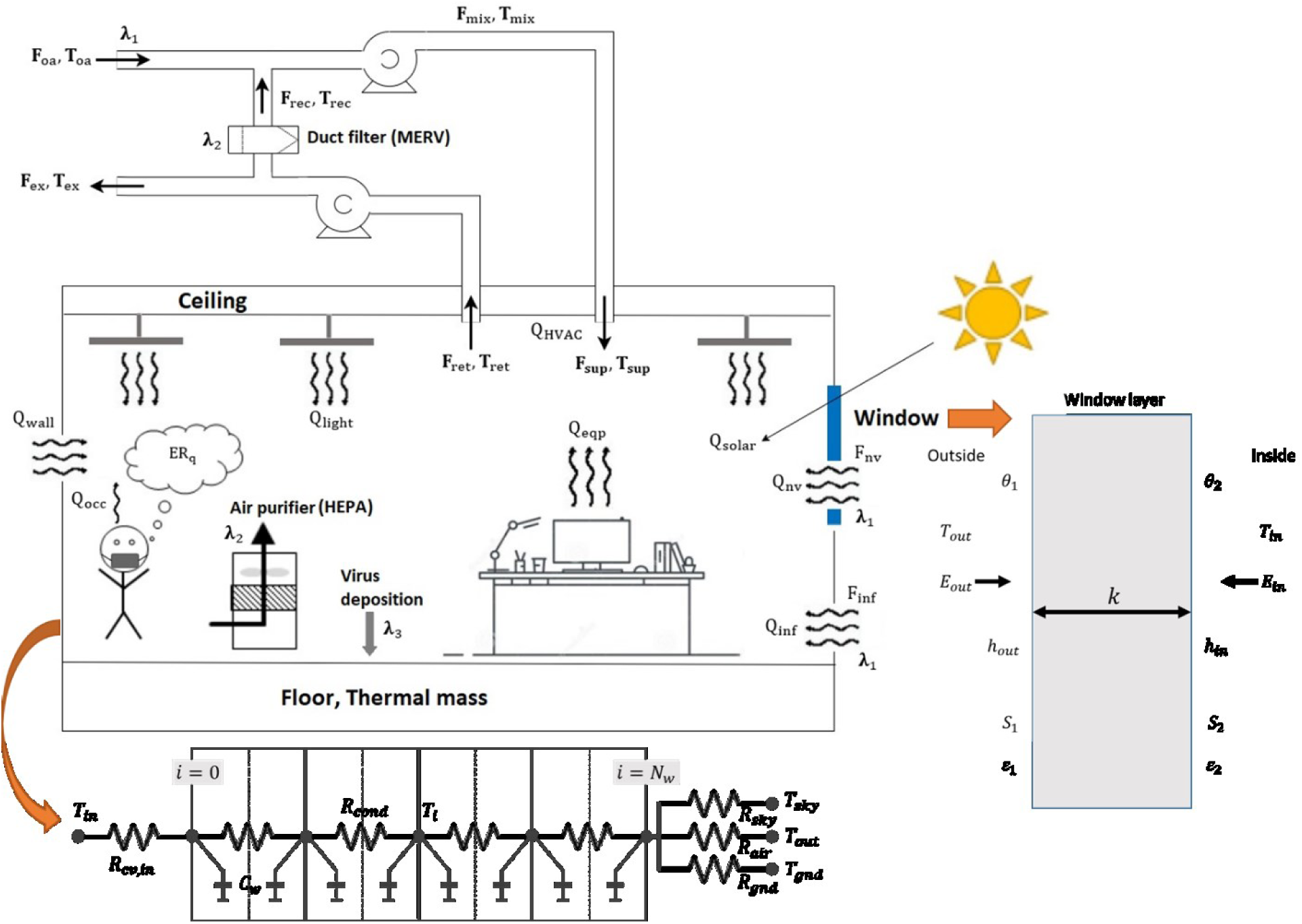
Schematic of the integrated CityRPI and CityBEM.

### 2.1. CityRPI for assessing aerosol infection risks and mitigation strategies

The aerosol infection risk calculation is based on the modified Wells-Riley formulation [30,31]. The following theory was based on the original work of the COVID-19 Aerosole Transmission Estimator” by Jiminez et al. (2020) while this study extended the work to various building types at a whole-city scale for North America, investigating various mitigation strategies and their energy impacts, which have not been addressed previously. The estimator is based on five assumptions: i) one infector in a space with a constant SARS-CoV-2 quanta generation rate, ii) zero initial quanta in the space, iii) latent period of the disease is longer than the time duration of the event, iv) indoor environment is well-mixed, and v) the infectious quanta removal is a first-order process. Two types of P.I. are calculated: conditional P.I. and absolute P.I. The conditional probability of infection (*PI*_*cond*_) calculates the probability of another person getting infected, under the condition that there are infectors in the room. It is calculated based on the assumed number of infected people inside the room, and the prevalence rate of the disease in the studied area is neglected in the calculation. The absolute probability of infection (*PI*_*abs*_) is calculated based on the *PI*_*cond*_ and considering the prevalence rate of the disease in a community/city/region.

### Conditional probability of infection

The conditional probability of infection (*PI*_*cond*_) is calculated by Eq. 1:

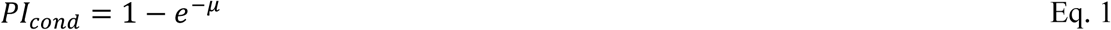

where *μ* is the number of quanta inhaled by a susceptible. Consider a susceptible in the room who inhales at rate B (*m*^3^/*h*) and is present for T hours. The expected number of quanta inhaled is calculated by Eq. 2:

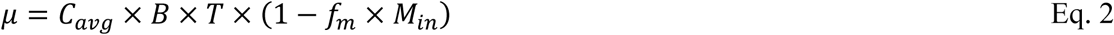

*C*_*avg*_ is the time-average quanta concentration (*q*/*m*^3^); *f*_*m*_ is the fraction of people in the room who wears the mask, and *M*_*in*_ is the inhalation mask efficiency. By solving the well-mixed material balance equation for the room (Eq. 3), the *C*_*avg*_ is calculated using Eq. 4.

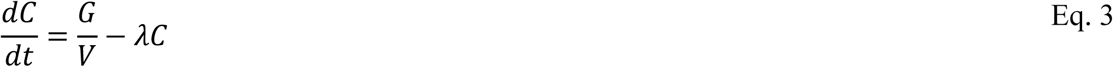

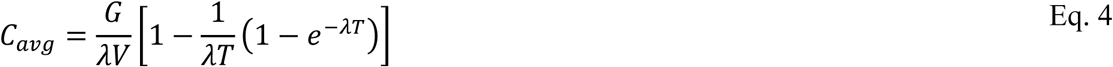

*V* is the volume of the room (*m*^3^); *λ* is the first-order loss rate coefficient for quanta (*h*^-1^); and *G* is the net quanta emission rate (*h*^-1^). *G* is calculated based on the number of infected people in the room (*N*_*inf*_), the fraction of people in the room with the mask (*f*_*m*_), exhalation mask efficiency (*M*_*ex*_), and quanta emission rate by one infected individual E. R._*q*_ (Eq. 5).

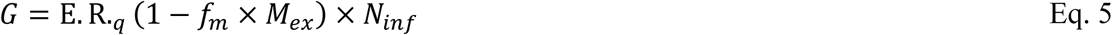

Buonanno et al. (2020a) calculated the quanta emission rate E. R._*q*_ of SARS-CoV-2 for different combinations of expiratory activities (oral breathing, speaking, and singing or loudly speaking) and activity levels (resting, light activity, and heavy exercise).

### First-order loss rate coefficient (λ)

The infectious quanta could be removed from room air by first-order processes reflecting several mechanisms: outdoor air ventilation *λ*_1_, filtration *λ*_2_, deposition on surfaces *λ*_3_, and airborne inactivation *λ*_4_.

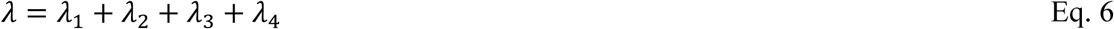

First, infectious quanta are removed with a first-order rate constant *λ*_1_ equal to the air change of outdoor air per hour (*h*^-1^) supplied to the room by the HVAC system or opening the windows. Second, the in-room air filtration using portable air purifiers and/or duct filters in HVAC systems can remove infectious quanta with a rate constant *λ*_2_. Third, infectious quanta are removed by gravitational settling with a first-order rate constant *λ*_3_. The deposition rate is calculated by [15] and is equal to 0.24 *h*^-1^. Finally, infectious quanta are inactivated with a first-order rate constant *λ*_4_. The quanta inactivation was evaluated based on the SARSCoV-2 half-life (1.1 h) and is equal to 0.63 *h*^-1^ [33].

### Absolute probability of infection

For estimation of infection risk in city-scale where the number of infected individuals in each building is not known, the prevalence of the disease in the community is used to estimate how many infected individuals may be present in the building, and the absolute probability of infection *PI*_*abs*_ is calculated based on estimated infected individuals [34].

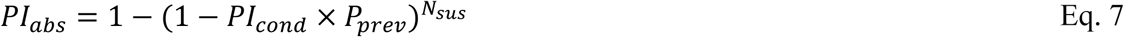

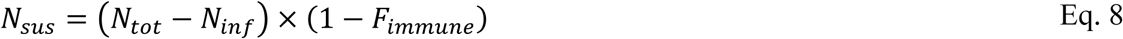

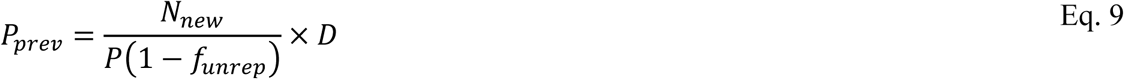

*P*_*prev*_ is the disease prevalence in the community that depends on the state of the pandemic in the region of study and the period of the disease. *N*_*new*_ and *P* are the number of daily new cases and the population of the studied region, respectively. *f*_*unrep*_ is the fraction of unreported cases. A study on ten diverse geographical sites in the U.S. shows that the estimated number of infections was much greater (6 to 24 times) than the number of reported cases in all sites [35]. *D* is the duration of the infectious period of SARS-CoV-2 [36]. *N*_*tot*_ and *N*_*sus*_ are the total number and number of susceptible people in the room, respectively. *F*_*immune*_ is the fraction of the population that has had the disease and has some immunity against it. It can be estimated using the total recovered cases in the region of study [35,37].

### 2.2. CityBEM for assessing urban building energy impacts

CityBEM [27,28] is an urban building energy model covering all essential heat and mass transfer mechanisms for the calculations of building heating/cooling loads, energy consumption, and indoor air and buildings’ surface temperature. The schematic of the models is shown in Fig. 1. CityBEM solves transient heat balance equations for indoor air, wall, floor, roof, thermal mass, and windows and calculates each element’s temperature. CityBEM models a Constant Air Volume (CAV) system for both heating and cooling. Heating/cooling energy consumption is estimated by calculating the building’s heatig/cooling load and modeling the HVAC system. The details for the CityBEM model are presented in Appendix A.

### 2.3. CityRPI archetype library

Airborne infection risk calculation using the equations of section 2.1 is a function of several buildings related parameters such as floor area, ceiling height, average stay time, the average age of occupants, occupant density, outdoor airflow rate, etc. Also, some buildings’ geometrical and non-geometrical properties are required for the calculation of buildings’ energy performance. Buildings’ geometry is obtained from open data sources such as OpenStreetMap (OSM) [38], Microsoft building footprint [39,40], and Google Earth (G.E.) API [41] as described by [28]. It is almost impossible to access other parameters for all buildings in a city. Parameters necessary for building energy performance calculation are obtained from an archetype library developed by Katal et al. (2019). We developed a new archetype library to estimate the necessary parameters for the P.I. calculation in this work. In this library, we classified buildings into twenty-nine different usage types. Then, we assigned the required parameters to each class. The usage type of buildings for the City of Montreal studied in this work is obtained from the shapefiles of boundaries of the property assessment units (PAU) [42].

We used several standards, codes, and references to collect the information for creating the archetype library. The number of occupants and outdoor ventilation rate are estimated based on ASHRAE Standard 62.1-Table 6.2.2.1 [26]. The table provides occupant density *D*_*occ*_ (#/100*m*^2^), outdoor airflow rate required per person *R*_*p*_ (*cfm*/*person*), and outdoor airflow rate required per unit area *R*_*a*_ (*cfm*/*ft*^2^) for different occupancy category (building type). Using these parameters and the floor area of the building *A*_*f*_ (*m*^2^), total number of occupants *N*_*tot*_ and air change of outdoor air per hour *ACH*_*vent*_ (*h*^-1^) is estimated by Eqs. 10 and 11.

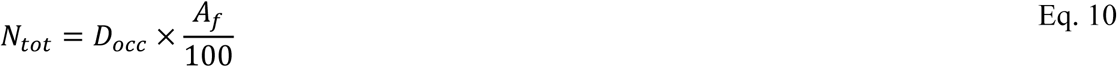

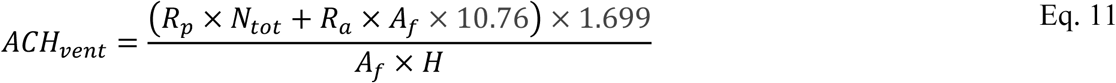

The susceptible person’s breath rate in the room is estimated using US EPA Exposure Factors Handbook (Chapter 6) (U.S. Environmental Protection Agency, 2011). The breath rate depends on age and activity level. A typical activity type and average age are defined for each building type to assign the buildings’ breath rate. The quanta emission rate by an infective person in the room depends on the activity level. We used the recommended quanta emission rate by Buonanno et al. (2020a, 2020b) and standard activity levels to estimate each building type’s quanta emission rate. The ceiling height and stay time are obtained based on the standard values of different building types. Therefore, in this work, we assumed that all buildings are designed and operating based on the available standards to calculate the indoor airborne infection risk and energy consumption. Table 2 shows some key input parameters of the archetype library assigned to each building type.

### 2.4. Interpolation of HRDPS weather data for buildings

Using local weather data for building energy simulation instead of nearby weather station data such as airports can significantly affect its accuracy [27,28]. In this work, mesoscale weather data provided by the High-Resolution Deterministic Prediction System (HRDPS) [44] is used to simulate all buildings in the City of Montreal. The spatial and temporal resolutions of the HRDPS model are 2.5 km and 1 hour, respectively. The Predictions are performed four times a day: 12 AM, 6 AM, 12 PM, and 6 PM. The forecasting horizon is 48 hours. HRDPS data are extracted and then interpolated on each building: First, the simulation area’s boundaries (Latitude and Longitude) and all HRDPS grid points located inside the domain are specified. In this work, Montreal City is selected for the simulation (Fig. 2). Gray circles show the HRDPS grid points. Then, atmospheric elements required for building energy simulation (outdoor air temperature, solar radiation, wind speed, wind direction, and dew point temperature) are extracted for all HRDPS grid points inside the domain. Finally, four HRDPS grid points around each building are selected, and weather data are interpolated on the center of the building based on the distance between the building center point and grid points [45].

**Fig. 2.**
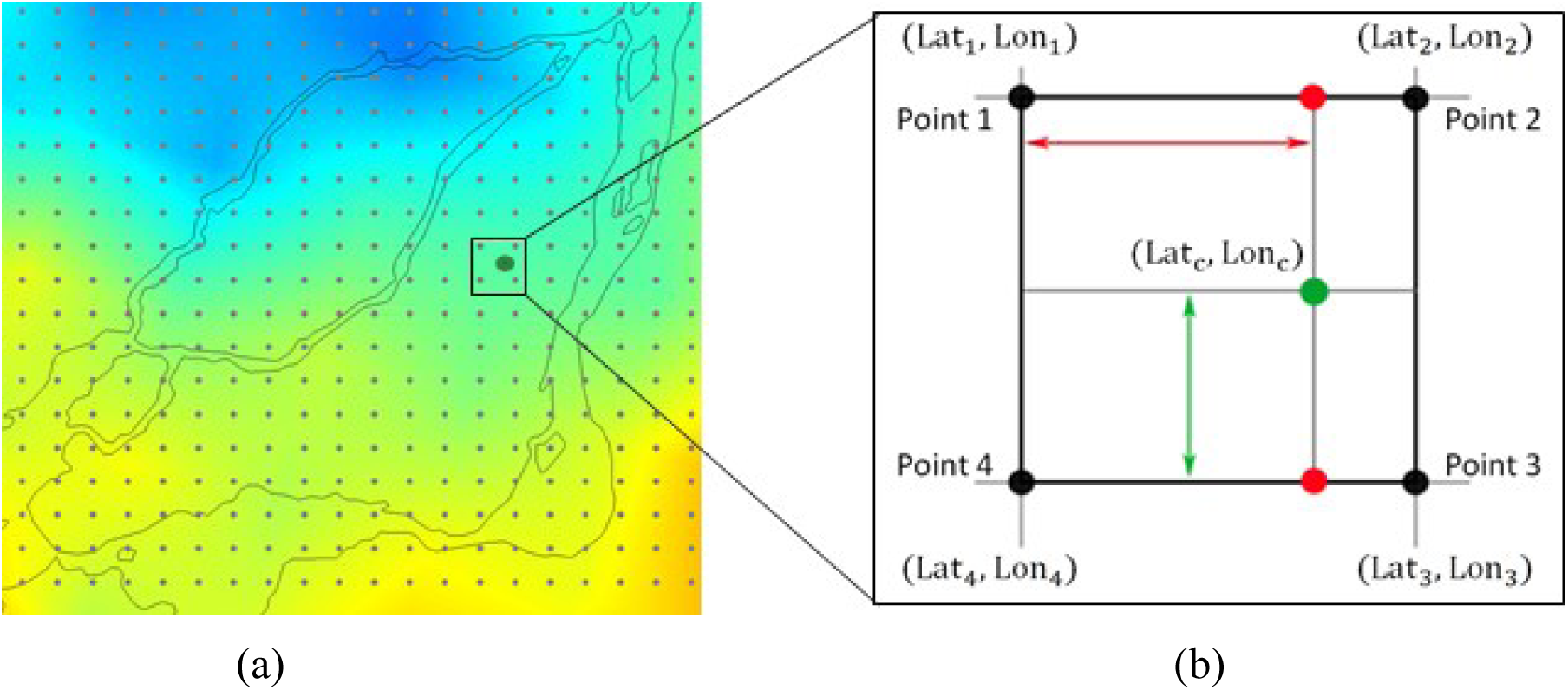
a) HRDPS weather data and grid points used for CityRPI simulation, b) Schematic of HRDPS data interpolation on the building center point.

### 2.5. CityRPI web application

To provide real-time data on airborne infection risk in North American cities, we developed an interactive website for the CityRPI model (https://concordia-cityrpi.web.app/). The purpose is to calculate the airborne infection risk in all buildings of a city and evaluate the impact of different mitigation measures on reducing the risk. The interactive website helps individuals find the best strategies for reducing the airborne infection risk in their buildings. The number of daily new infected cases in U.S. and Canadian cities are obtained from a GitHub page created by the New York Times [46] and a GIS Hub provided by Esri Canada [47]. We calculate a baseline infection risk for each building based on the normal condition, then calculate the reduced probability of infection (RPI) using seven different mitigation measures. By moving the mouse pointer on a building, an instant pop-up window shows the effectiveness of strategies sorted from the most to the least effective strategy. Fig. 3a shows an example pop-up window for a school building in Montreal on January 5, 2021. For this building, based on the archetype library’s information, wearing a face mask and adding 30% more outdoor air are the most and least effective strategies, respectively. The user can click on the building and then change the building information, including the number of occupants, floor area, ceiling height, ventilation and recirculation rate, exposure time, and lockdown and reopening dates (Fig. 3b). The results section provides a more detailed analysis and historical trend of the infection risk since the beginning of the pandemic.

**Fig. 3.**
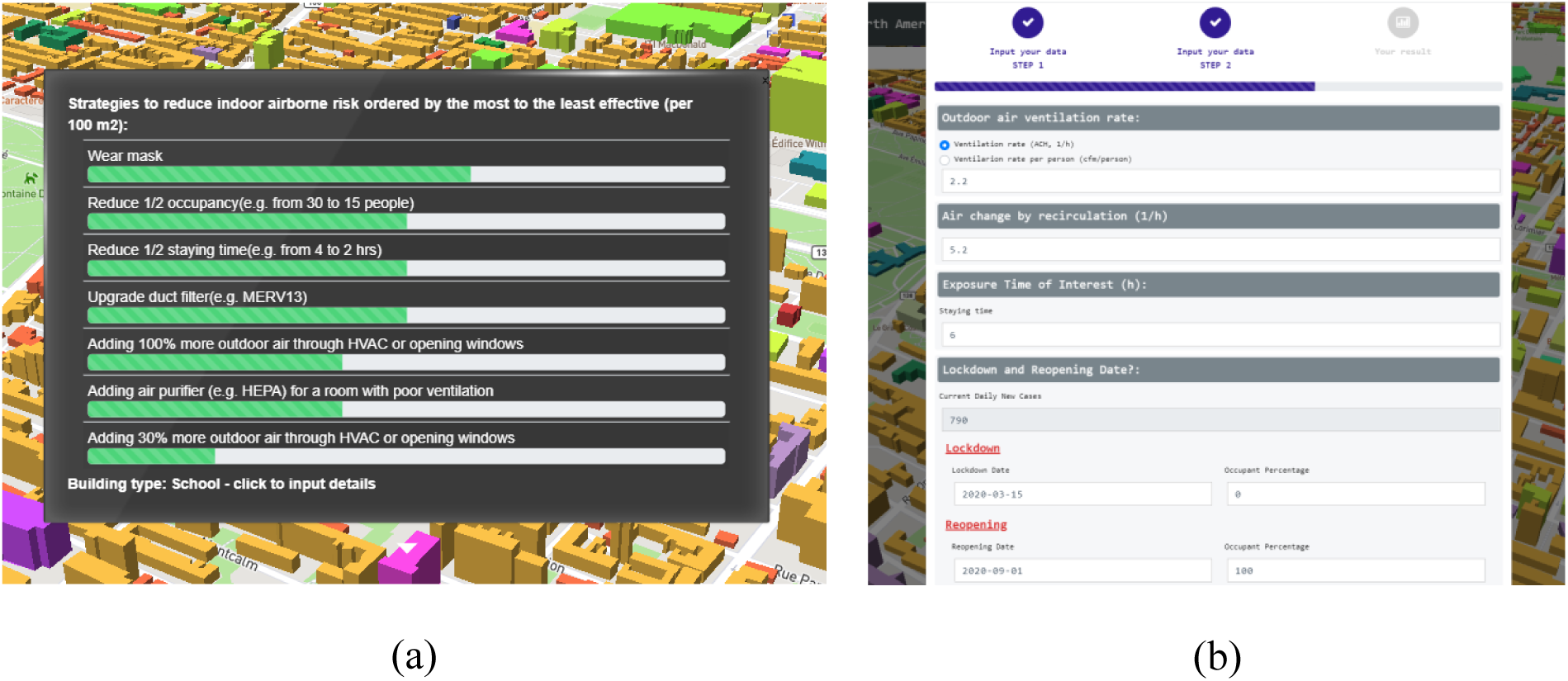
CityRPI website: a) Instant pop-up window for the effectiveness of different strategies, b) Input window for building information details and results display.

## 3. Verification and Validation

In this section, the CityRPI model’s accuracy is investigated by performing separate validations on the aerosol infection risk model and indoor air temperature simulation.

### 3.1. Verification of CityRPI aerosol infection risk calculation

To verify the airborne infection risk model, we modeled a real super spreading event (SSE) in Skagit Valley, Washington, USA. One symptomatic case attended the Skagit Valley Chorale (SVC) weekly rehearsal on March 10, 2020. Among the 61 attendees at the event, 53 cases were subsequently infected. Detailed analysis of evidence and reports by Miller et al. (2020) shows that respiratory aerosols’ airborne transmission was the leading mode of transmission in this event. Miller et al. (2020) estimated the average quanta emission rate based on the available information and calculated the conditional P.I. for different scenarios. Jimenez (2020) extended the previous work by Miller et al. (2020). He developed the COVID-19 Aerosol Transmission Estimator (COVID-19 ATE) to calculate conditional and absolute airborne infection risk in several indoor environments. He evaluated the disease prevalence rate at the Skagit Valley and calculated the event’s absolute P.I. In this section. We modeled the SVC event by the CityRPI and compared the conditional and absolute P.I. with the results by Miller et al. (2020) and Jimenez (2020). Table 3 shows the simulation settings of the SVC event. Conditional and absolute probabilities of infections are calculated as a function of loss rate *λ* and results are shown in Fig. 4. Infection risks calculated by CityRPI are the same as Miller et al. and COVID-19 ATE, which shows that the airborne infection risk calculation by CityRPI is verified.

**Fig. 4.**
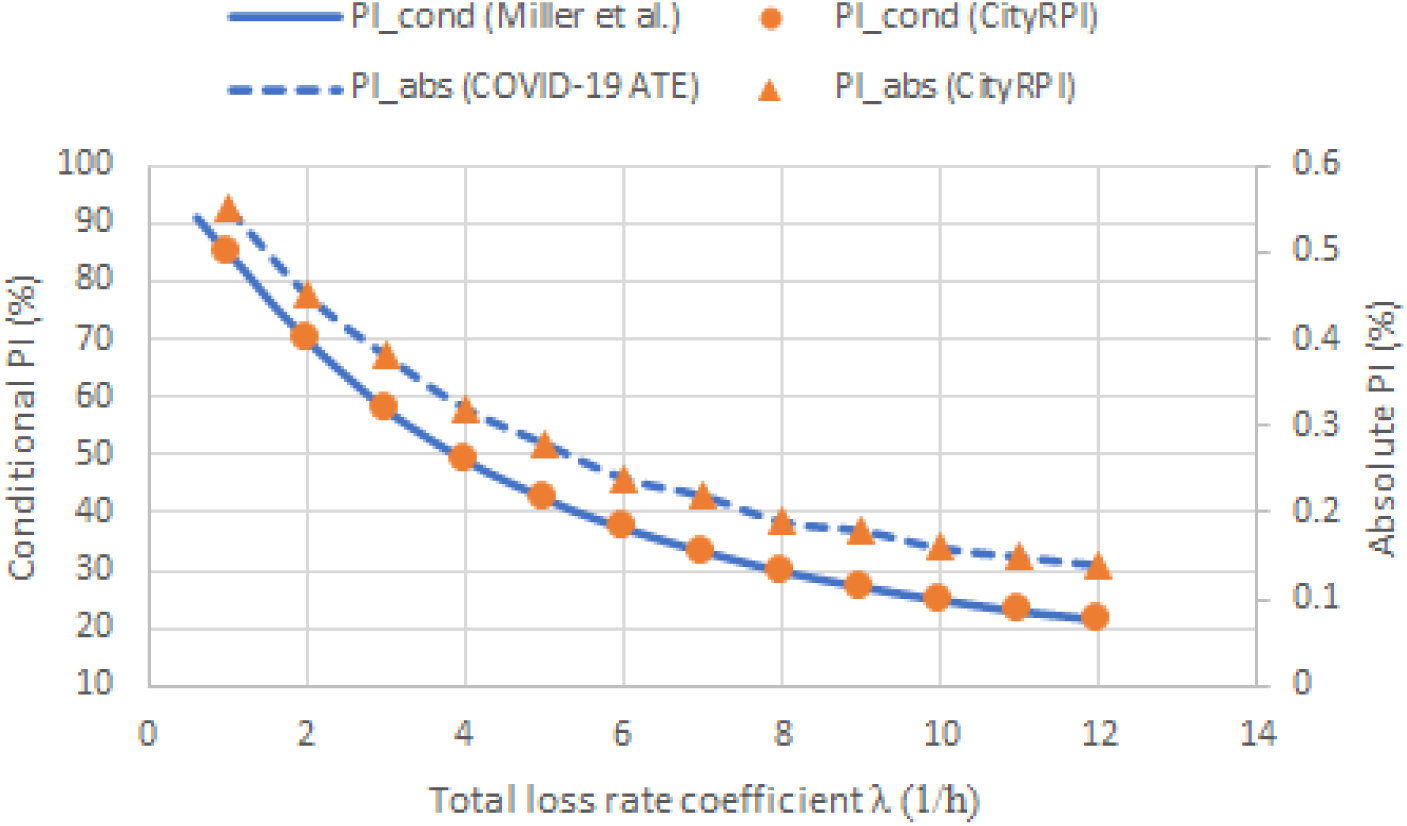
Comparison of *PI*_*cond*_ and *PI*_*abs*_ at different loss rate coefficients for SVC super spreading event calculated by CityRPI, COVID-19 ATE from Jimenez (2020), and by Miller et al. (2020).

### 3.2. Validation of CityBEM energy calculation

The accuracy of the CityBEM for the calculation of building energy performance is already investigated by comparing the annual and hourly electricity consumption with corresponding measurement data [27,28]. In this work, for the validation of indoor air temperature results calculated by CityRPI, Montreal City is simulated from June 24-29, 2020. We compared the calculated indoor air temperature of two school buildings with measurement data provided by sensors installed in the rooms. Buildings are not equipped with an Air conditioning (A.C.) system, or the A.C. systems were not operating at the study time. Therefore, the indoor air temperature is directly affected by the cooling load components, and an accurate calculation of indoor air temperature could indicate the accuracy of building energy load estimation. Figs. 5a and 5b show the schools’ aerial view map and weather data (outdoor air temperature and solar radiation) used for the simulation, respectively. According to Fig. 5b, the studied days were quite hot and sunny. In the absence of A.C. systems in the buildings, indoor air temperature is much higher than cooling-setpoint temperature.

**Fig. 5.**
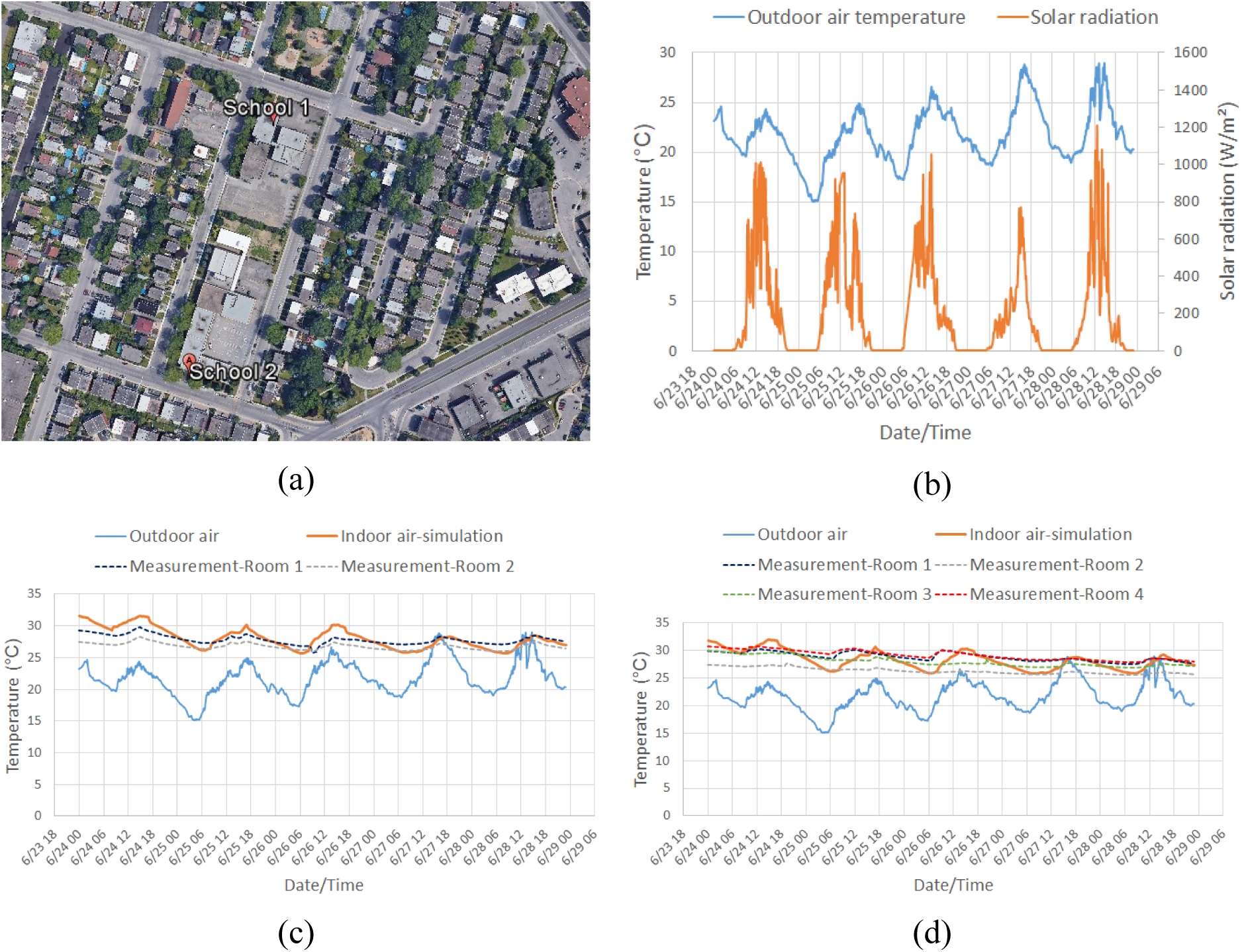
Validation of indoor air temperature calculated by CityRPI: a) aerial view map,b) weather data of simulation period, c) comparison of simulation and measurement data of school 1, d) comparison of simulation and measurement data of school 2.

School 1 is a two-story building constructed in 1953. Two indoor sensors are installed in two classrooms on the first floor. School 2 is a two-story building constructed in 1958 with four indoor sensors to measure indoor air temperature. There is no A.C. system in the classrooms, so the HVAC model is disabled in CityRPI simulation. Figs. 5c and 5d compare the indoor air temperature calculated by CityRPI with all sensors data. The accuracy of the result is acceptable considering the uncertainties of the input parameters used for CityRPI simulation and the simplified single-zone model used for the whole building. The RMSE between the average temperature of classrooms and simulation results of schools 1 and 2 are 1.8 °C and 1.6 °C, respectively.

## 4. Case study

According to Equations 1-9, the indoor aerosol infection risk is a function of several parameters. Some of them, such as the number of occupants, ventilation rate, stay time, and breathing rate, depend on the building usage type. Therefore, infection risk and the effectiveness of mitigation measures can change by building type. In this section, we modeled the Montreal City by CityRPI and studied the impact of six mitigation strategies on buildings’ indoor aerosol infection risk. Increasing the outdoor air ventilation rate by opening the windows or more outdoor air from the HVAC intakes can improve the room ventilation condition and reduce infection risk. But it can also significantly increase building energy consumption in the winter. Therefore, it is essential to find the most effective strategy considering both the reduced probability of infection and the building’s energy consumption. For this purpose, we conducted the simulation over Montreal from February 12-21, 2020, which was the coldest period of winter 2019, and the effectiveness of three mitigation strategies for improving the indoor ventilation condition (more outdoor air, upgrading duct filter, portable air cleaner) are compared.

### Geometry and input data

The simulation area that covers Montreal City is shown in Fig. 2. We generated the 3D model of buildings using Microsoft building footprint data and Google Earth building height information [28]. The total number of buildings is 358,295. Building type and year of construction are obtained from the Montreal PAU shapefile [42]. COVID-19 statistical data in Montreal, including daily new cases, totally confirmed cases, total deaths, and recovered cases, are provided by Esri Canada [47]. Other parameters used to calculate infection risk are obtained from the archetype library (Table 2).

**Table 1.** Buildings archetype library for calculation of airborne infection risk.

| Building | H<sub>f</sub>(m) | R<sub>p</sub><br>(cfm/person) | R<sub>a</sub><br>(cfm/ft<sup>2</sup>) | Occupant<br>density<br>(#/100m<sup>2</sup>) | Stay<br>time (h) | E. R.<sub>q</sub><br>(q/h) |
| --- | --- | --- | --- | --- | --- | --- |
| Apartment | 3.3 | 5 | 0.06 | 4 | 10 | 5.70 |
| Assembly | 5 | 5 | 0.06 | 150 | 3 | 15.95 |
| Bank | 3.2 | 7.5 | 0.06 | 15 | 1 | 6.85 |
| Cottage | 2.5 | 5 | 0.06 | 3 | 10 | 5.70 |
| Courthouse | 4.8 | 5 | 0.06 | 70 | 3 | 6.85 |
| Daycare | 2.5 | 10 | 0.18 | 25 | 6 | 15.95 |
| Hospital | 3 | 7.5 | 0.18 | 20 | 2 | 6.85 |
| Hotel | 3.6 | 5 | 0.06 | 10 | 2 | 6.85 |
| House | 2.5 | 5 | 0.06 | 3 | 10 | 5.70 |
| Industrial | 9 | 10 | 0.18 | 7 | 4 | 15.95 |
| Library | 4.5 | 5 | 0.12 | 10 | 3 | 6.85 |
| Museum | 6.5 | 7.5 | 0.06 | 40 | 3 | 15.95 |
| Nursing home | 2.5 | 5 | 0.12 | 25 | 2 | 5.70 |
| Office | 4 | 5 | 0.06 | 5 | 4 | 6.85 |
| Parking | 2 | 5 | 0.06 | 25 | 0.5 | 15.95 |
| Personal service | 3 | 7.5 | 0.06 | 25 | 1 | 6.85 |
| Public service | 2.5 | 5 | 0.06 | 30 | 1 | 15.95 |
| Restaurant | 3 | 7.5 | 0.18 | 70 | 1.5 | 6.85 |
| Retail store | 4.5 | 7.5 | 0.06 | 40 | 1 | 15.95 |
| Retirement house | 2.5 | 5 | 0.06 | 10 | 2 | 5.70 |
| School | 3 | 5 | 0.12 | 35 | 6 | 6.85 |
| Shared house | 2.5 | 5 | 0.06 | 10 | 2 | 5.70 |
| Shopping center | 2.4 | 7.5 | 0.06 | 40 | 2 | 15.95 |
| Sport center | 6 | 20 | 0.18 | 7 | 2 | 38.30 |
| Student house | 2.5 | 5 | 0.06 | 10 | 2 | 5.70 |
| Studio | 2.5 | 5 | 0.12 | 10 | 4 | 6.85 |
| Transport | 2.5 | 7.5 | 0.06 | 100 | 0.5 | 6.85 |
| University | 3 | 10 | 0.18 | 25 | 2 | 6.85 |
| Warehouse | 6 | 10 | 0.06 | 4 | 1 | 15.95 |

**Table 2.** Parameters used for the simulation of SVC super spreading event

| Parameter | Symbol | Unit | Value |
| --- | --- | --- | --- |
| Volume | $V$ | $(m^3)$ | 810 |
| Duration of event | $T$ | hour | 2.5 |
| Volumetric breathing rate | $B$ | $(m^3/h)$ | 1.0 |
| Quanta emission rate | $E. R_q$ | $quanta/h$ | 970 |
| Fraction of population with immunity | $F_{immune}$ | - | 0 |
| Fraction of people with mask | $f_m$ | - | 0 |
| Total number of people in the room | $N_{tot}$ | - | 61 |
| Number of infective people | $N_{inf}$ | - | 1 |
| Disease prevalence rate | $P_{prev}$ | % | 0.011 |

## Results

In this section, first, we study the conditional and absolute P.I. of buildings at baseline conditions (e.g., minimum design ventilation rate by default). We compare the infection risk between different building types to find the most vulnerable buildings against the airborne infection risk of COVID-19. We also study the daily variation of *PI*_*abs*_ to investigate its change with the prevalence rate of the disease. Then, to reduce the infection risk in buildings, we designed several scenarios using different mitigation measures. We applied the mitigation measures to all buildings in the city to find the most effective strategies for each building type. Some strategies also affect building energy consumption. Therefore, we calculated the increased peak demand for all buildings and identified the recommended strategy for risk reduction and energy consumption.

### Probability of infection and prevalence rate

To study P.I. variation with the city’s prevalence rate, we calculated all buildings’ historical P.I. from the start of the pandemic in Montreal until January 6, 2021. The first wave of the COVID-19 pandemic started on February 25, 2020, and the complete lockdown measures began on March 18. By reducing the number of daily cases, the reopening gradually started from May 2 with the reopening of schools and daycares around January 1. The second wave of pandemic begun around the middle of September and the number of daily new cases is still rising. The Quebec government reimposed a partial lockdown on December 25 that will be continued until February 8. The red dot-line in Fig. 6 shows the weekly prevalence rate of the COVID-19 in

**Fig. 6.**
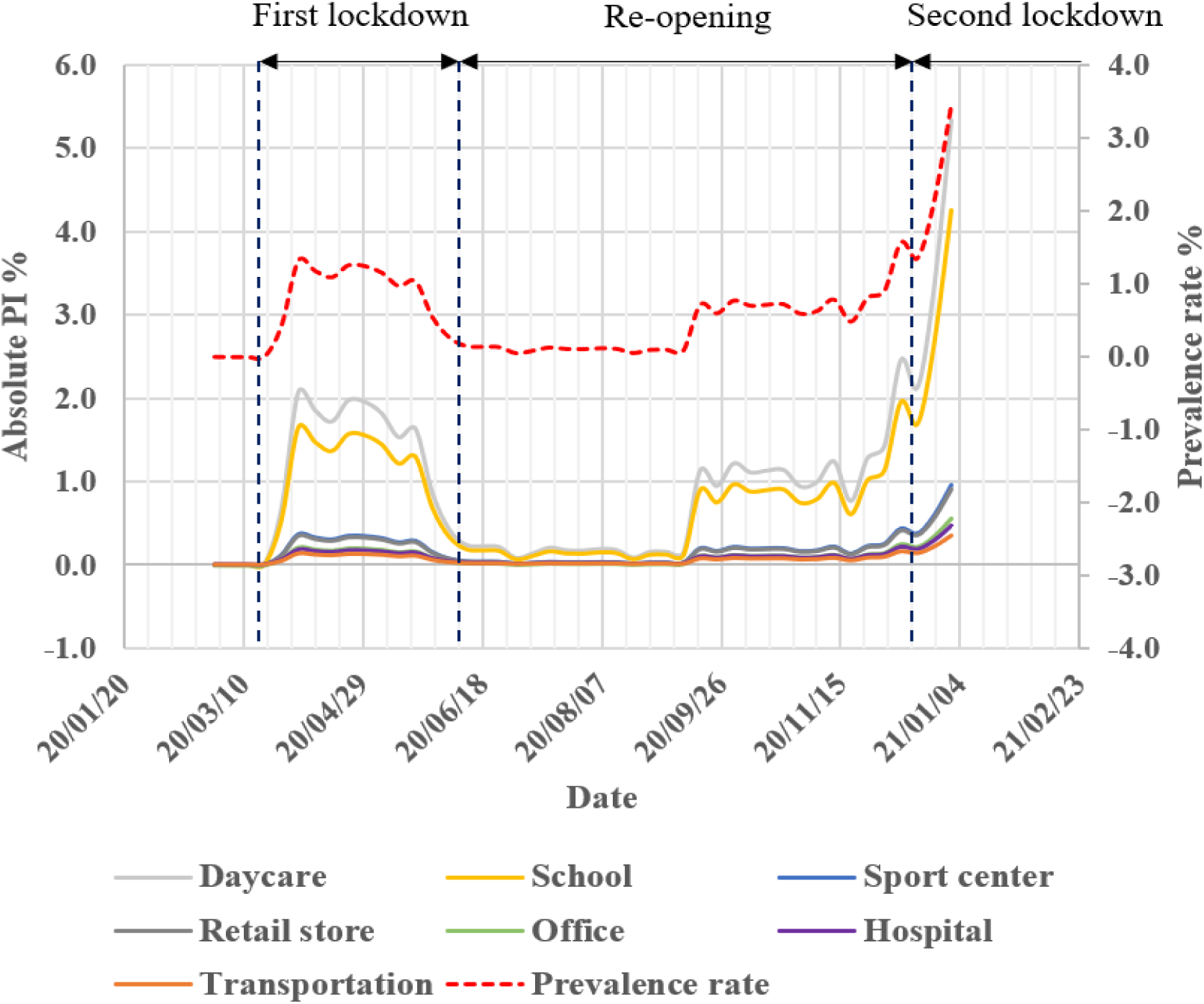
The historical prevalence rate of COVID-19 in Montreal and *PI*_*abs*_ of seven selected building types.

Montreal. The highest prevalence rate is around 4%, which means that one person could be infected among 25 persons averaged over the whole population. It includes both symptomatic and asymptomatic cases. The historical *PI*_*abs*_ of seven building types are plotted and compared in Fig. 6. We calculated the *PI*_*abs*_ of all 29 building types, but to compare the variation of relative *PI*_*abs*_ with time, the result is plotted for seven buildings with the most usage and importance during the pandemic. We did not plot the *PI*_*cond*_ of buildings because it is not a function of the prevalence rate and is constant with time. According to Fig. 6, even though the *PI*_*abs*_ varies with the prevalence rate but the relative infection risk of buildings is the same; for example, daycare remains with the high *PI*_*abs*_ among selected buildings.

To compare the P.I. between all twenty-nine building types, we calculated the conditional and absolute P.I. of buildings on the worst day (May 3, 2020) with the largest daily new cases and prevalence rate. The number of daily new cases, total number of confirmed cases, total number of deaths, and the total number of recovered patients were 1652, 16251, 1365, and 9123. The prevalence rate of the disease was around 4.03%. Fig. 6 shows the weekly prevalence rate, and this specific day is not plotted in this figure, but as it can be seen, the number of daily new cases is still increasing during the second wave and arrives in close to the worst case.

### City-scale probability of infection of buildings

Figure 7a shows the *PI*_*abs*_, *PI*_*cond*_, and the number of buildings in each type per total number of buildings. This figure provides information to find the most vulnerable buildings against the COVID-19 airborne transmission. Buildings are sorted based on the *PI*_*abs*_ value. The daycare, classroom, studio, sports center, industrial building, and office room are six buildings with the largest *PI*_*cond*_. All these buildings except the sports center are buildings with long exposure time (> 4 hours). Sports center shows high *PI*_*cond*_ because of the increased quanta emission rate from heavy activities. Therefore, exposure time and quanta emission rate are two dominant parameters for the aerosol transmissions indoors.

**Fig. 7.**
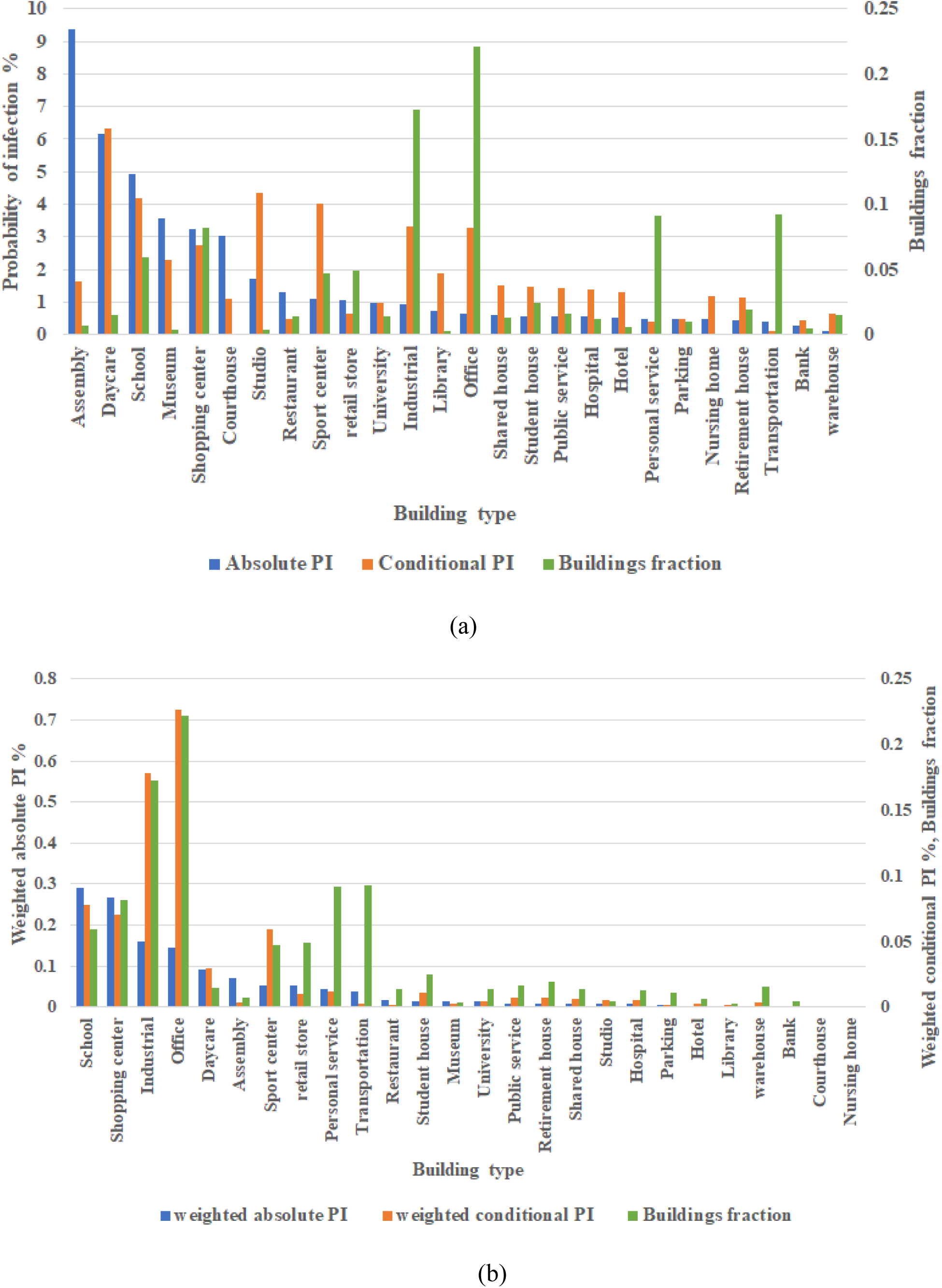
a) Conditional and absolute P.I. of all building types on the worst day of the pandemic, b) weighted conditional and absolute P.I., and building type fraction on the worst day.

The *PI*_*abs*_ is a function of the *PI*_*cond*_, the number of susceptibles, and the prevalence rate. In this case, the prevalence rate is the same for all buildings. The number of susceptible people depends on the number of occupants in the room. Unlike the conditional P.I., an assembly building shows the highest *PI*_*abs*_ because of the largest occupant density and susceptible people. Daycare, school, museum, shopping center, and courthouse are other buildings with large *PI*_*abs*_, due to high occupant density and *PI*_*cond*_.

The number of buildings per type is another important parameter when comparing different buildings. One building type can have a large *PI*_*abs*_, but if there are few buildings of this type in a city or urban area, it may not affect the city-scale risk. To consider its impact, we calculated the weighted probability of infection, which is the probability of infection multiplied by building type fraction, *PI* × *N*_*b,type*_/*N*_*b,total*_, which *N*_*b,type*_ is the number of buildings with a specific type and *N*_*b,total*_ is the total number of studied buildings. Most of the city buildings are residential buildings, but because our focus is on public buildings, residential buildings are removed from the analysis. Fig. 7b shows the weighted conditional and absolute P.I. The school buildings offer the largest weighted *PI*_*abs*_ because of large *PI*_*abs*_ and building fraction. Shopping centers, industrial buildings, offices, and daycares are the next vulnerable buildings. Regarding the weighted *PI*_*cond*_, which is “person to person” transmission without considering the prevalence rate, office, industrial building, school, shopping center, sports center, and daycare are the most vulnerable buildings. The office building was the 6^th^ vulnerable building considering only the *PI*_*cond*_, however, it is the most vulnerable building at the city scale because of the large number of office buildings in the city. Therefore, to reduce the city’s airborne infection risk, it is essential to focus on the buildings with the largest weighted P.I.

As mentioned previously, airborne infection risk depends on building properties. The *PI*_*abs*_ in Fig. 7b changes from 9.4% for an assembly room to 0.4% for a warehouse. Each person should determine the tolerable infection risk for himself/herself based on the situation, but some researchers have suggested a 0.1 % infection risk as to the acceptable risk level [34]. Results show that the probability of infection in all buildings on the worst day of the pandemic is higher than the tolerable risk level. Therefore, the existing standards, such as the required ventilation rate recommended by ASHRAE Standard 62.1., occupant density, and stay time, cannot provide a safe indoor environment. One or several mitigation strategies should be applied to reduce the COVID-19 airborne infection risk for indoor spaces.

### Reduced Probability of Infection and prevalence rate

Six different mitigation measures are studied for their impact on all building airborne infection risk, including wearing a face mask, reducing the stay time, reducing the number of people, using more outdoor air ventilation, upgrading the duct filter in the HVAC system, and using a portable air cleaner in the room. The baseline measure for comparison is no mask usage, minimum outdoor air ventilation, no duct filter, and air cleaner, full occupancy, and standard stay time.

Before analyzing the impact of scenarios on all buildings, it is crucial to study the RPI change with the prevalence rate. For this purpose, we calculated the classroom daily RPI from the beginning of the pandemic until January 6, 2021 (Fig. 8). We selected the school building because it is the most vulnerable building based on the Fig. 7b. Results show that all strategies’ effectiveness is almost constant with time and is independent of the prevalence rate. For a classroom, wearing a face mask by all occupants is the most effective strategy with 64% RPI. The next effective strategies are upgrading the duct filter to the MERV-13 filter, using a portable air cleaner with 480 CFM flow rate, half stay time, and half occupancy with 50%-58% RPI. Doubling outdoor air ventilation rate seems the least effective strategy with only 31% RPI.

**Fig. 8.**
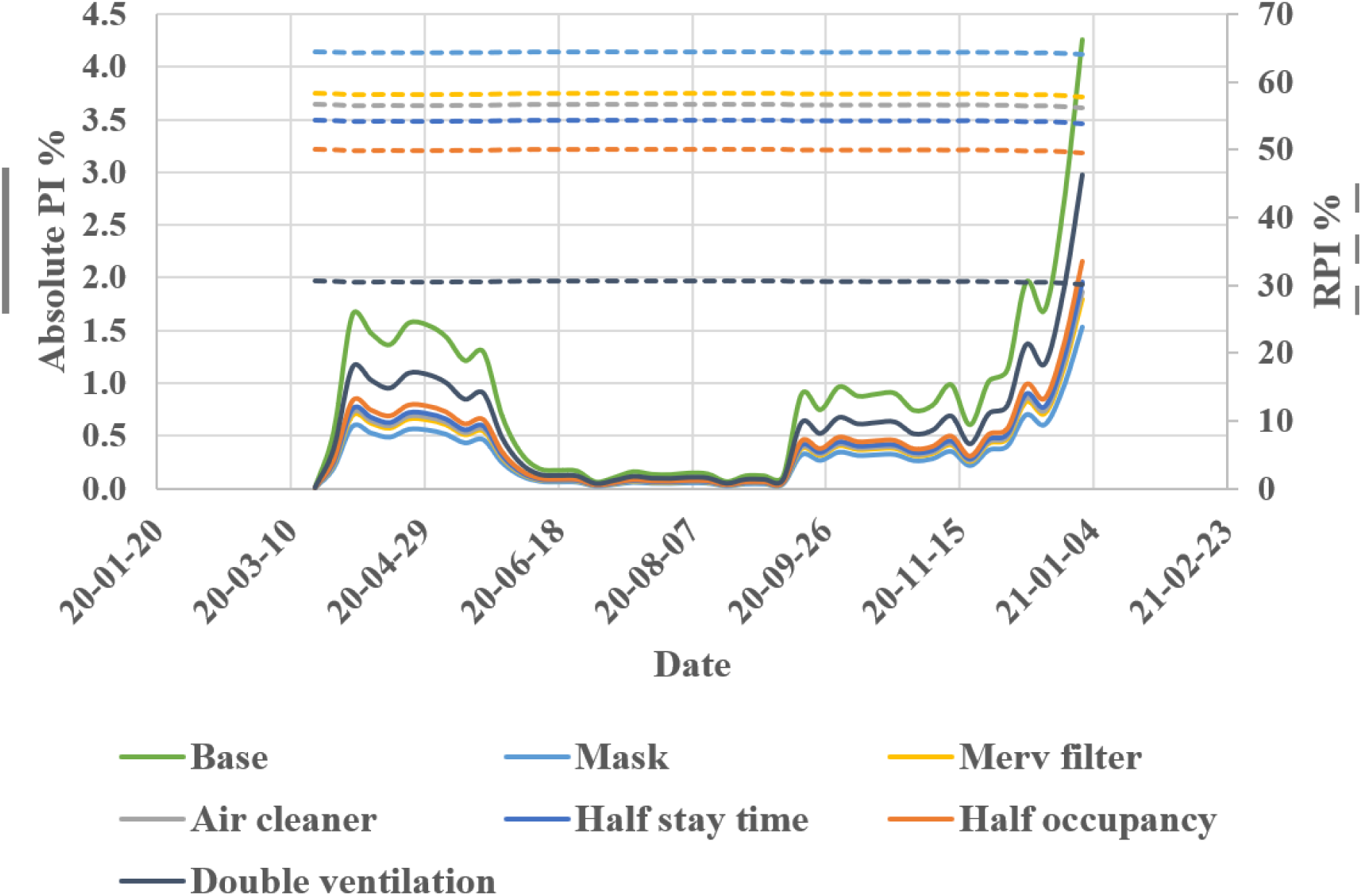
Variation of P.I. and RPI of school classroom for mitigation measures.

It should be noted that the duct filter RPI is the median of all school buildings in the city because the HVAC system airflow rate depends on the design condition and building properties. A study by Curtius et al. (2020) on using portable air cleaners in the classrooms show that a total air exchange rate of 5.7 h^-1^ reduces the airborne transmission by 80%, the equivalent air exchange rate of air cleaner modeled in this work is around 2.7 h^-1,^ and the reduced infection risk is 53%. By using more air cleaners in the room, we can get more reduction. The conditional P.I. is not plotted in this figure because it is independent of the prevalence rate.

According to Fig. 8, the relative effectiveness, RPI, of all strategies seems independent of the prevalence rate. Therefore, to study the impact of mitigation measures on all building infection risk reduction, we calculated the RPI on the worst pandemic day. Energy impact is another important factor that must be considered. Some of the strategies can increase building energy consumption while reducing the infection risk. For example, more outdoor air ventilation rate in the winter needs more pre-heating and thus more energy. Upgrading to duct filters with higher efficiency may increase fan power consumption, and a portable air cleaner also adds extra electricity usage. The number of daily cases in Montreal is rising during the winter; therefore, we calculated the increased building peak energy demand for the coldest period of winter 2019 by assuming the winter of 2020 would have similar weather.

### Effectiveness of mitigation measures and energy impacts

Figure 9 represents the reduced conditional and absolute P.I.s and peak energy demands of all building types obtained by different mitigation measures. Strategies are sorted based on the effectiveness of reducing the *PI*_*abs*_ of each building type. Comparing the reduced percentage of conditional and absolute infection risks shows that the reduction is around the same range for both P.I.s except for the half occupancy strategy. The *PI*_*cond*_ is “person-to-person” airborne transmission; therefore, the number of occupants does not impact the *PI*_*cond*_, while the number of susceptible people is directly related to the total number of occupants in the room and reducing the number of occupants will reduce the *PI*_*abs*_.

**Fig. 9.**
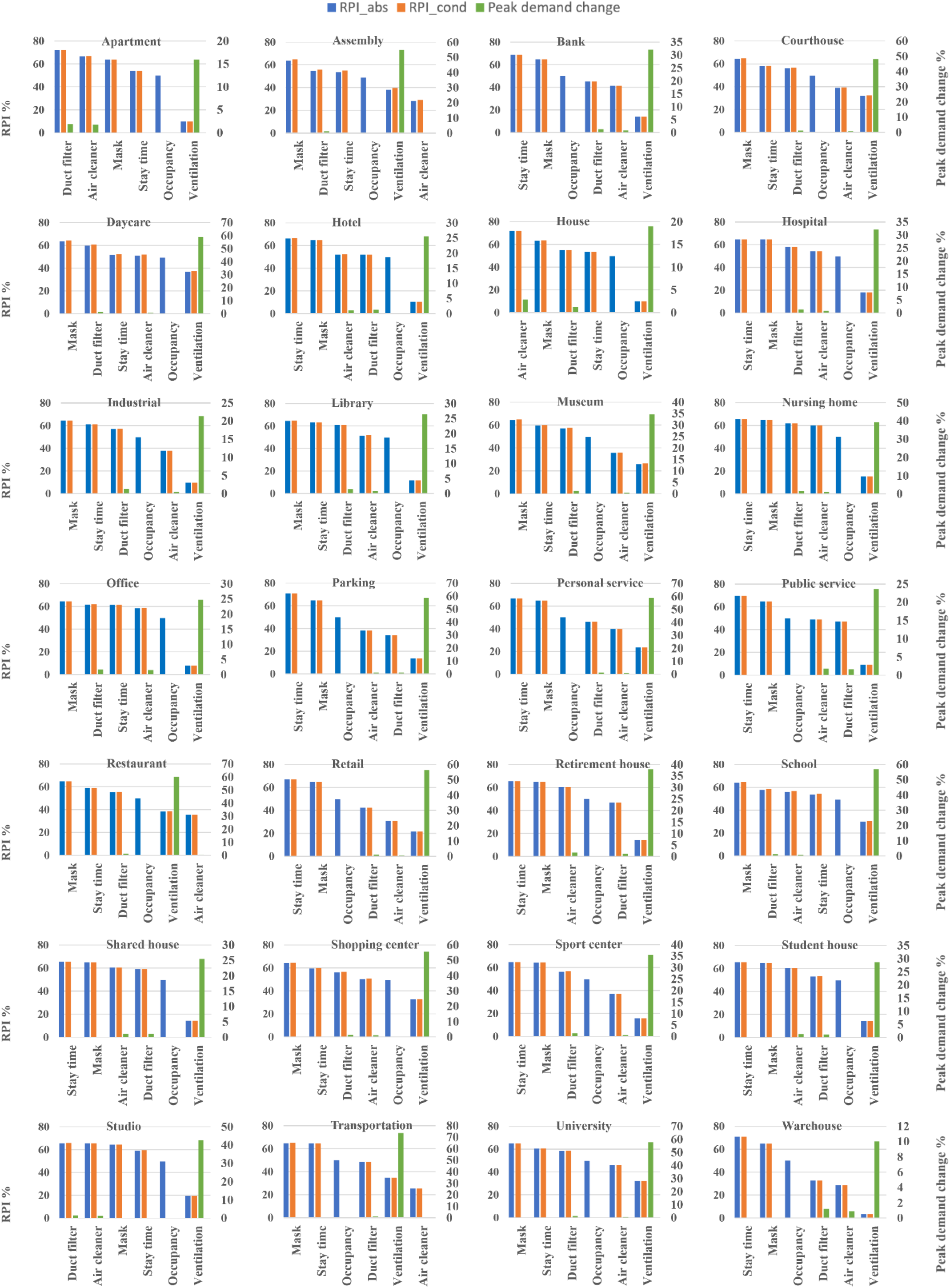
Conditional and absolute RPI and peak load change of strategies by building type.

Wearing a face mask or half stay time is the most effective strategy for reducing the infection risk in most buildings, except studio and residential buildings. The duct filter or using a portable air cleaner can be the most effective strategy in these buildings. On the other hand, double outdoor air ventilation rate is the least effective strategy in almost all the buildings except the assembly room, restaurant, and transportation environment. Using a portable air cleaner is the least effective strategy in these three building types. Proper outdoor air ventilation by opening the windows or air intakes can reduce COVID-19 aerosol particle concentration and minimize infection risk. The existing ASHRAE recommended minimum outdoor air ventilation rate for indoor air quality purposes is not enough for providing a safe indoor space regarding the airborne transmission of COVID-19 aerosol respiratory. To reduce the infection risk, ASHRAE recommended increasing the outdoor air and reducing the HVAC system recirculation air. Fig. 8 shows that a 100% increase in outdoor air ventilation rate is not as effective as the other strategies.

The distribution of buildings by usage type is plotted in Fig. 10a. We plotted the *RPI*_*abs*_ of all buildings in the City downtown for double outdoor air ventilation rate and wearing a face mask in Figs. 10b and 10c. This region includes diverse building types and covers almost all building types studied in this work. The RPI range for double outdoor ventilation rate is between 0.1%- 35% but for wearing a face mask is always larger than 60%. Our analysis shows that four times outdoor air ventilation rate could provide a similar level reduction of infection risk as wearing a face mask in many cases, consistent with the result presented by Dai and Zhao (2020).

**Fig. 10.**
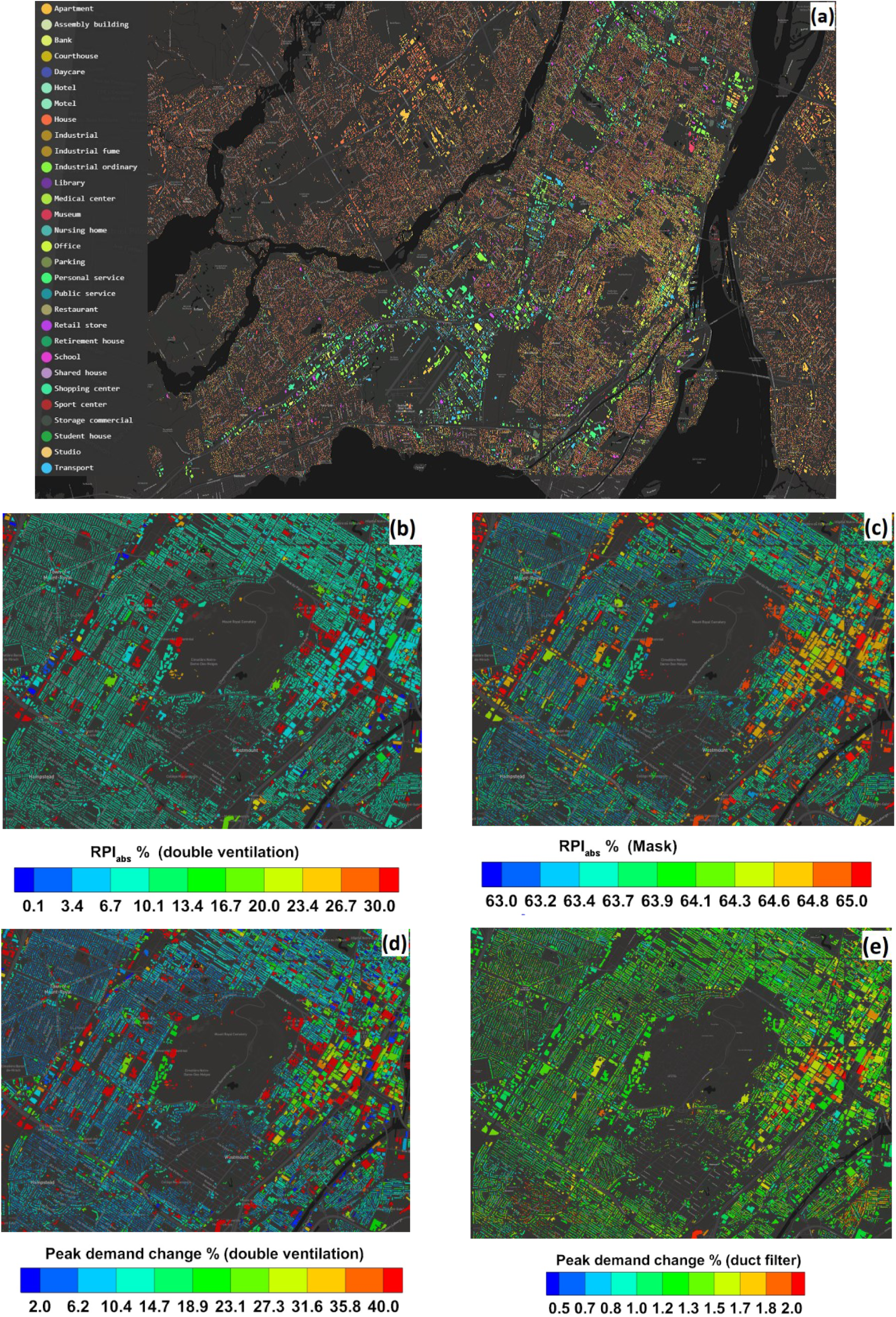
a) building distribution by type, b, c) RPI by double outdoor air ventilation rate and wearing a face mask, d,e) Peak energy demand change by double outdoor air ventilation and upgrading the duct filter.

The increased peak energy demand for double outdoor air ventilation varies between 10%-60% depending on the building type. At the same time, it is less than 2% for upgrading duct filters (MERV 13) or using a portable air cleaner, and other strategies do not have an energy penalty. The results are plotted in Figs. 10d and 10e. The energy penalty using double outdoor air ventilation rate is between 2%-70% for all buildings while it is less than 2% for upgrading the duct filter. Considering both reduced infection risk and increased peak demand, we may conclude that double outdoor air ventilation rate is the least effective strategy. To get the same RPI as other strategies such as wearing a face mask or half stay time, we need to further increase the outdoor air ventilation rate while unavoidably increasing the heat demand. As a result, this could build excessive pressures on the electricity grid and increase the chance of power outages in the city.

In conclusion, all studied measures can contribute to reducing the indoor aerosol infection risk of COVID-19. The effectiveness of these measures depends on building types and properties. Several strategies can be used together for more reduction in the risk and providing a safer indoor environment. We recommend wearing a face mask or reducing half of the stay time as the first and most effective strategy in all buildings based on the presented results. Reducing the stay time and occupant number can significantly reduce the risk, especially in crowded spaces and for buildings where people often have to linger for a while. Regarding improving ventilation conditions to reduce infection risk, upgrading the duct filter of HVAC systems and using a portable air cleaner are more effective than merely increasing outdoor air ventilation rates. The former measures also have a lower energy penalty than the increased outdoor ventilation rate.

## 5. Conclusion and Future Work

This study developed the CityRPI model to calculate the airborne transmission of COVID-19 in a city. We developed an archetype library based on standards and references to estimate different parameters for calculating building infection risk. We modeled Montreal City and calculated the infection risk of all buildings in the city, and evaluated the impact of six mitigation strategies on reducing infection risk. Some strategies impact the energy consumption of a building, especially in the winter. We integrated CityRPI with the CityBEM model to calculate all building’s peak energy demand. The main conclusions are as follows:

1. On the day with the highest infection rate, the probability of infection in all buildings was higher than the tolerable infection risk. It shows that buildings’ standard operating conditions, such as the minimum ventilation rate recommended by ASHRAE, occupants’ density, and stay time, were not enough for the occupants’ safety. Stay time and quanta emission rate are the two dominant factors regarding the infection risk in buildings.
2. The weighted probability of infection shows that, on a whole city scale, school buildings are the most vulnerable in the city currently, considering the absolute P.I. Offices are the most vulnerable buildings, considering the conditional P.I., which is the “person-to-person” airborne transmission.
3. The mitigation study shows that wearing a face mask and half stay time are the most effective strategies for most buildings. Double outdoor air ventilation rate is the least effective strategy in many buildings, and the corresponding RPI is much lower than other strategies.
4. Double outdoor ventilation rate also significantly increases building peak heating demand in the winter. Therefore, this strategy is not as effective as other strategies considering the infection risk and electricity consumption in the winter.
5. The CityRPI website provides separate infection risk calculations and most to the least effective strategies for each building in the city. A user can change the input parameters and properties of his/her building to improve the calculation accuracy and get a more detailed analysis.

The calculation of infection risk involves many input parameters, and the results could be subject to the uncertainties of these parameters. As future work, a sensitivity analysis of the infection risk model becomes necessary to quantify these uncertainties’ impacts on the results. It will help to determine which parameters are dominant and improve the accuracy of the estimated values. Meanwhile, the current infection risk model could underestimate the risk, especially for an event with many occupants and for a high prevalence rate in a city because the conditional probability is based on the scenario that there is only one infector present in a room. With more people and higher prevalence, the chance of more than one infectors in the same room could become high, which could be considered in a future study.

## Data Availability

The data in this manuscript are available from the CityRPI website.

https://concordia-cityrpi.web.app/

## Acknowledgments

The research is supported by the Natural Sciences and Engineering Research Council (NSERC) of Canada through the Discovery Grants Program [#RGPIN-2018-06734] and the Advancing Climate Change Science in Canada Program [#ACCPJ 535986-18]. The authors acknowledged the valuable discussions with Dr. Chanjuan Sun at the University of Shanghai for Science and Technology, China, Dr. John Zhai, and Dr. Shelly Miller at the University of Colorado, Boulder, USA.

